# ChatIBD: design, safeguards, and early international use of a guideline-grounded generative AI tool for inflammatory bowel disease professionals

**DOI:** 10.64898/2026.05.06.26352526

**Authors:** Cher Shiong Chuah, Beatriz Gros, Nikolas Plevris

## Abstract

**Objectives:** To describe the design, operational safeguards, and early use of ChatIBD, a specialty-specific generative AI platform for inflammatory bowel disease (IBD) healthcare professionals, during its first 6 months of live deployment.

**Methods:** ChatIBD (www.chatibd.com) is an online question-answering platform that uses retrieval-augmented generation over a curated corpus of IBD guidelines. Queries undergo hybrid semantic and keyword retrieval with query expansion and reranking, and the model is instructed to answer only from retrieved material and return linked citations. Safeguards include fixed medication dosing information from European Medicines Agency (EMA), user feedback capture, and clinician review of flagged outputs. We performed a descriptive service evaluation of aggregated, de-identified platform metrics collected between 1 October 2025 and 1 April 2026. Primary outcomes were uptake and repeat engagement; secondary outcomes were geographic and language distribution, message domain and intent, dosing-card activation, and user feedback events.

**Results:** During the study period, ChatIBD registered 913 users and processed 7,222 user messages across 3,855 conversations. Of 913 registered users, 684 (74.9%) sent at least one message and 349 (38.2%) met the pre-specified active-use threshold of three or more messages. Activity was recorded across 69 countries and 28 languages, with the highest message volumes from the United Kingdom (27.1%) and Spain (12.3%). Median daily message volume was 35.5 (IQR 20 to 52), and 85.1% of messages were submitted on weekdays. Medication-related queries accounted for the largest use domain, while guideline synthesis was the most frequent inferred intent. Sixteen explicit feedback events were recorded, including one negative rating that triggered clinician review and system changes.

**Conclusions:** ChatIBD showed early international uptake and repeat use as a specialty-specific, guideline-grounded generative AI tool for IBD professionals, deployed with a set of practical operational safeguards. These usage findings do not establish clinical accuracy, safety, or effectiveness. Formal validation is in progress.

**What is already known on this topic:** General-purpose large language models are increasingly being used informally by clinicians, but concerns remain about hallucinated content, unverifiable recommendations, and poor traceability to specialty-specific sources.

**What this study adds:** This study describes the early deployment of ChatIBD, a specialty-specific retrieval-grounded generative AI tool for IBD professionals, and the safeguards used in its live operation.

**How this study might affect research, practice or policy:** Early evaluations of live specialist AI tools may help guide governance, implementation, and validation. Uptake alone is not evidence of effectiveness, but it can help shape priorities for subsequent studies.

## Introduction

Inflammatory bowel disease (IBD), comprising Crohn’s disease (CD) and ulcerative colitis (UC), is a growing global health challenge, with rising incidence in newly industrialised regions and compounding prevalence in established western healthcare systems.(1) At the same time, IBD management has become increasingly knowledge-intensive. Therapeutic options have expanded, and clinicians must navigate a rapidly evolving evidence base. For time-pressured clinicians, keeping pace is increasingly difficult.(2)

Large language models are attracting growing interest across medicine, and clinicians are increasingly experimenting with artificial intelligence (AI) tools for information retrieval, summarisation, and documentation support.(3) However, the use of general-purpose systems for specialist clinical questions raises important concerns, particularly around hallucinated or unsupported content.(4–6)

One response is to develop specialty-specific systems that constrain model behaviour through retrieval over curated source documents and embed additional safeguards around the interaction.(7,8) Rather than treating the model as a source of clinical knowledge, this approach uses it as an interface layer over an explicitly selected evidence base.

ChatIBD (www.chatibd.com) was developed with this rationale. It is a web-based generative AI tool designed to answer IBD-related clinician questions using retrieval-augmented generation (RAG) over curated guideline content, with cited responses and additional safeguards for higher-risk query types.

Here, we detail the implementation and report a descriptive service evaluation of ChatIBD during its first 6 months of live deployment, focusing on system design, safeguards, and early use patterns rather than clinical effectiveness. This study provides an early real-world account of a specialty-specific generative AI tool in gastroenterology and may inform similar efforts in other clinical domains.

## Methods

### Study design

We undertook a descriptive service evaluation of ChatIBD during its first 6 months of public deployment. Analysis was restricted to aggregated, de-identified platform-level metrics generated during routine service operation. The study period ran from 1 October 2025 to 1 April 2026. This descriptive implementation and early-use study is reported with reference to the DECIDE-AI reporting guideline for the early-stage clinical evaluation of decision-support systems driven by artificial intelligence (Supplementary Table 2). (9)

### Development approach and intended use

Development of ChatIBD began in July 2025, with public beta deployment on 1 October 2025. The platform was conceived and developed by three practising IBD gastroenterologists. The clinical scope and requirements were defined jointly by the three co-founders. The corresponding author led product design and technical implementation, while the other authors led content curation, user engagement, and validation planning.

ChatIBD was designed as a clinician-facing guideline retrieval and summarisation tool for healthcare professionals and researchers with a professional interest in IBD. Its intended functions were to identify and summarise recommendations from selected guidelines, compare recommendations across sources, and provide rapid access to curated medication dosing information. It was not designed to diagnose, prescribe, triage, monitor patients, make autonomous clinical decisions, or replace review of the underlying source material.

Registration was open to any user in either guest or authenticated mode, with no formal eligibility screening, and no target sample size was set. No structured user training was provided; familiarisation relied on in-application onboarding, disclaimer text, and the cited-source interface.

### Technical architecture

ChatIBD was implemented as a web-based application using Next.js and the Vercel AI SDK (Figure 1). At public launch, responses were generated using OpenAI GPT-4.1 On 18 November 2025, the production model was changed to GPT-5.1 following clinician review of approximately 50 representative queries. GPT-5.1 was selected because it was judged to produce better responses and mitigated limitations identified from early production operations.

**Figure 1.**
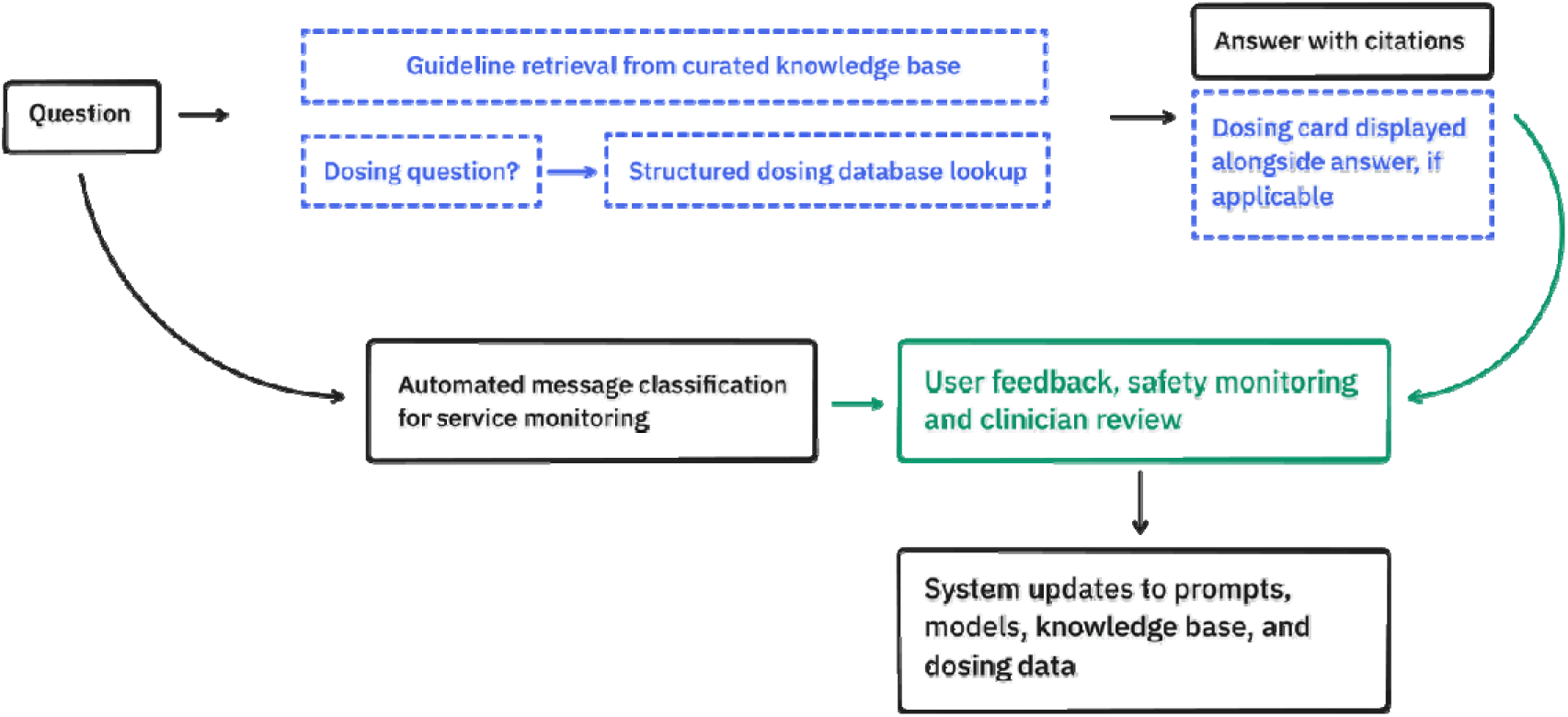
ChatIBD platform architecture and safety workflow.

During the study period, model inference was provided through the OpenAI and xAI application programming interface under commercial terms that excluded submitted data from use in model training. The model and prompt configuration associated with each generated response were recorded to support traceability. Changes to models, prompts, retrieval behaviour, and source content were maintained through version control and internal change logs.

### Knowledge-base selection and processing

The primary knowledge base consisted of prospectively selected IBD guidelines and related guidance documents chosen by the three co-founders. Eligible documents were required to be clinically relevant to IBD, attributable to a recognised professional, academic, regulatory, or medicines organisation, date-stamped, available in a stable and citable format, and suitable for lawful indexing.

Priority was given to current UK-relevant guidance, major international IBD society guidelines, regulatory medicines information, and paediatric-specific guidance. At launch, the corpus contained 32 documents published from 2019 onwards. This increased to 35 during the study period following the addition of updated documents following informal community feedback. The complete corpus used during the study period is provided in Supplementary Table 1.

Source documents were ingested into a managed vector store (OpenAI). Each document was segmented into passages of approximately 800 tokens with 400-token overlap and indexed using OpenAI text-embedding-3-large embeddings. Extracted content and retrieval outputs were reviewed by sampling during development to identify parsing, formatting, or content-selection errors.

### Retrieval and response generation

At query time, the user message, together with the preceding conversation where needed to resolve context, was submitted to the managed vector store. The vector store performed hybrid retrieval combining semantic (embedding-based) similarity with lexical keyword matching, with internal query rewriting, expansion, and reranking. The 15 highest-ranked passages were returned as context for response generation.

The system prompt instructed the model to answer only from the retrieved material, cite the supporting sources, and state clearly when relevant information could not be identified. Citations were rendered in the user interface as links to the corresponding source document (Figure 2A). For non-English questions, the response was generated in the language used by the user, while the indexed knowledge base was English.

**Figure 2.**
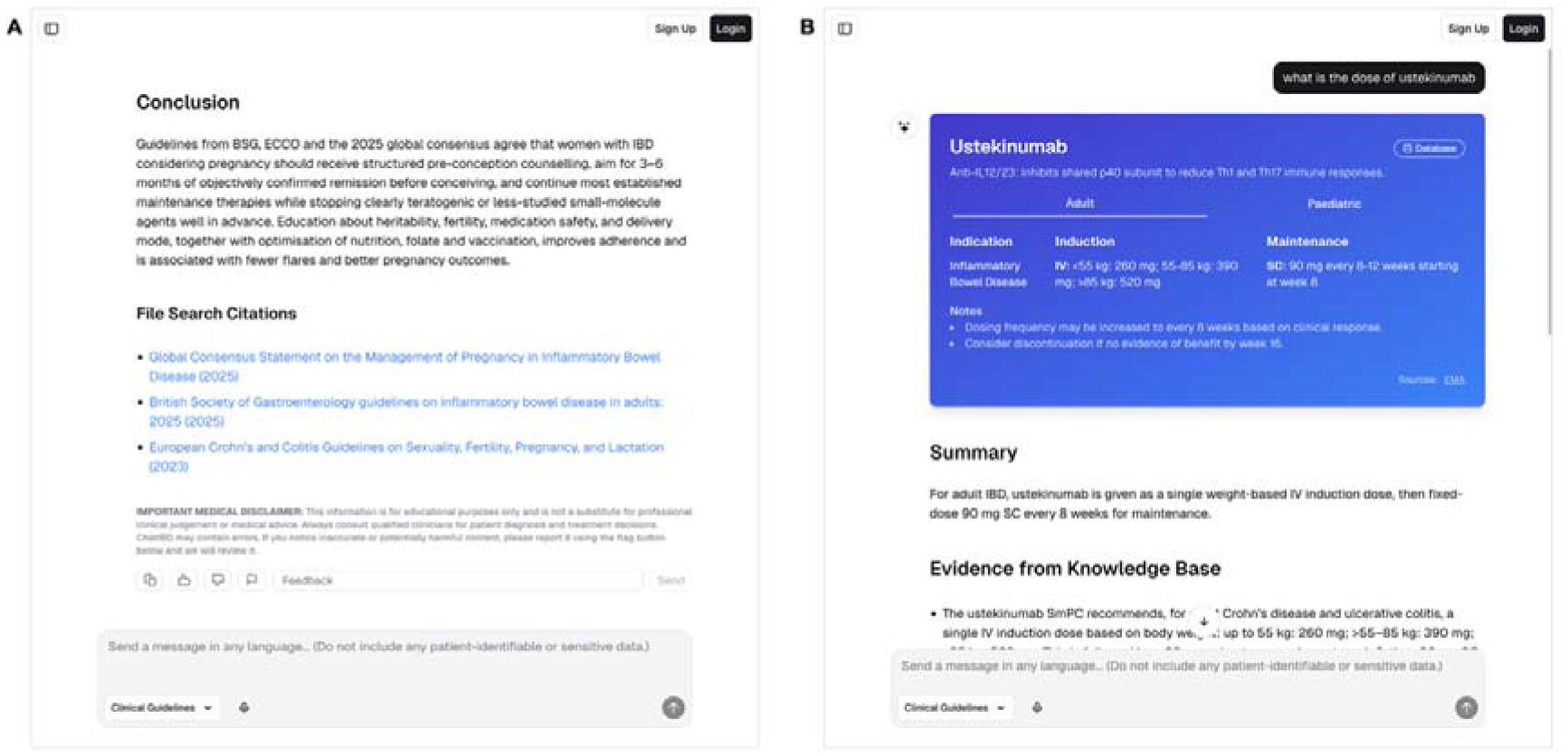
Example ChatIBD user interface. **A:** standard response showing grounded answer generation with source citations, disclaimer text, and user feedback controls. **B:** dose-specific query showing the dosing card displayed alongside the narrative response.

### Pre-deployment testing

Before the public beta launch, ChatIBD underwent a nine-week formative testing phase from 29 July to 30 September 2025. During this period, 553 test queries were submitted across 28 user accounts and 340 conversations.

The founding development team submitted 489 queries, representing 88.4% of all pre-deployment queries, across four accounts. The remaining 64 queries, representing 11.6%, were contributed by 24 invited testers, predominantly gastroenterologists based in the United Kingdom, Spain, and Latin America.

Queries were submitted mainly in English (86.3%) and Spanish (12.7%). Testing covered the principal domains of IBD practice. The most frequent domains were pharmacotherapy and medication dosing (35.4%), reproduction and lactation (12.5%), infection and immunisation (9.6%), education and disease-score interpretation (7.8%), comorbidities and special populations (6.7%), surveillance and monitoring (6.5%), perioperative and procedural care (5.6%), and diagnostics and laboratory interpretation (4.9%).

Outputs were reviewed during development for clinical relevance, clarity, citation support, and potentially unsafe interpretation. Identified issues informed iterative changes to prompts, retrieval behaviour, source content, and the user interface. No patient-identifiable information was entered during testing. This formative testing supported system development and was not intended to constitute formal clinical validation.

### Structured dosing database

Queries concerning medication dosing, schedules, or treatment protocols were analysed using a model-based classifier (grok-4-fast-reasoning) that identified listed IBD medications. When a recognised medication was detected, the platform displayed a dosing card alongside the narrative response (Figure 2B).

The structured dosing database was manually extracted and curated by the corresponding authors from public European Medicines Agency information and relevant professional guidance. Each medication was represented using structured fields including drug class, brand-name aliases, source information, adult and paediatric use, clinical indication, induction and maintenance treatment, route of administration, dose, and additional monitoring or safety notes.

The language model was used only to identify the medication referred to in the query. The content of the dosing card was retrieved deterministically from the version-controlled structured dosing database and was not generated by the language model. Where a medication was not recognised or no corresponding structured record was available, no dosing card was displayed.

### Operational safeguards and feedback review

An overview of operational safeguards is described in Table 1. Potential patient-identifiable information was addressed through instructions within the main system prompt and through reminder text displayed as a placeholder in the message input field. Where a message appeared to contain direct identifiers, such as a patient name or date of birth, the model was instructed to decline the request and ask the user to remove identifying information before resubmission. This was a prompt-based safeguard rather than a separately validated identifier-detection system.

**Table 1.** Operational safeguards and risk-mitigation features incorporated into ChatIBD.

| **Safeguard** | **Risk addressed** | **Implementation** |
| --- | --- | --- |
| Curated specialty-specific knowledge base | Generic or off-topic responses from unrestricted general-purpose AI | Responses were generated over a prospectively selected corpus of IBD guideline and guidance documents chosen by the co-founders, all practising IBD gastroenterologists |
| Retrieval-augmented generation with visible citations | Unsupported free-form generation and poor traceability of recommendations | User queries underwent retrieval of relevant source passages, and the model was instructed to answer only from supplied context and return linked citations |
| Patient-identifiable information screening | Entry of identifiable patient information | Messages appearing to contain direct identifiers prompted a model-generated request to remove identifying information before resubmission. This was a prompt-based safeguard rather than a validated identifier-detection control. |
| Structured dosing database | Higher-risk errors in medication dose, route, timing, or regimen | Dose-specific medication queries triggered a dosing card linked to a maintained medication dataset derived from public EMA information and displayed alongside the narrative response |
| User feedback capture | Problematic outputs going undetected after deployment | Users could rate responses or flag them as potentially inaccurate or clinically concerning |
| Flagged-output review and iterative system update | Recurrent errors persisting in a live system | Flagged outputs were reviewed by a clinician, categorised, and used to guide changes to prompts, models, or source content where needed |

The platform captured explicit user feedback on generated outputs. Users could rate responses and flag outputs considered inaccurate or clinically concerning. Flagged interactions entered a clinician-led review process in which the original question, retrieved evidence, generated response, and citations were examined. Review findings could result in changes to the system prompt, model, retrieval process, source corpus, structured dosing database, or interface. Revised behaviour was retested before deployment. During the study period, this workflow was led by the corresponding author without independent adjudication.

### Data governance, privacy, and security

The platform could be accessed without registration in a guest mode subject to a shared rate limit, or through an authenticated account managed by a third-party identity provider; administrative accounts supported platform operation. Conversation data were stored in a managed PostgreSQL database hosted in the United Kingdom (Amazon Web Services London region, eu-west-2), with encryption in transit and at rest. Application hosting and edge delivery were provided by Vercel, product analytics by PostHog, and error monitoring by Sentry; model inference during the study period was provided by OpenAI and xAI. All processors operated under data-processing terms, and submitted data were excluded from model training. Automated bot traffic was mitigated using the hosting provider’s bot-detection and blocking.

### Change management and auditability

All application code, system prompts, and knowledge-base configuration were maintained under version control (Git). In addition, each stored message recorded references to immutable model-version and prompt-version records, capturing the provider, model name, model configuration, and the exact prompt content and fingerprint used to generate that response. This provided a per-message audit trail from which the precise model and prompt configuration applied to any interaction during the study period could be reconstructed. Substantive changes to the system were additionally recorded in a publicly accessible changelog, providing transparency to users regarding updates to models, prompts, and source content. Changes in usage, feedback, or response behaviour following each modification were not formally assessed.

### Subsequent development

Platform behaviour was not static after the study period. Following the observation window, the retrieval pipeline was migrated from the managed vector store to a self-hosted corpus to enable sentence-level citation provenance. The production model was updated to improve latency, cost, and response quality, and model inference was migrated from the OpenAI and xAI APIs to Microsoft Foundry to standardise inference billing. These changes post-date the metrics reported here and are described to make clear that the configuration evaluated in this study represents a defined point-in-time deployment.

### Usage metrics and descriptive analysis

Metrics included cumulative registrations, message counts, conversation counts, active use, country and language metadata, optional self-reported professional background, dosing-card activation, feedback events, and temporal activity patterns. Metrics were extracted directly from the platform database.

A conversation was defined using the platform’s native chat-thread identifier, and a message as a discrete message event recorded within that thread. Active use was defined a priori as submission of three or more messages during the observation period. This threshold was selected as a pragmatic indicator of repeat engagement.

Country was inferred from internet-protocol-based geolocation metadata provided at the network edge (Vercel), and language from automated message-level language tagging. Professional background was derived from an optional self-reported user profile. Profile information was requested only after users had submitted three or more messages, and completion remained optional. Professional-background data were therefore available only for a subset of more engaged users and were not assumed to represent the full user population.

Continuous variables are reported using medians and interquartile ranges, and categorical variables using counts and percentages. No inferential statistical testing was performed.

### Outcomes

The primary outcomes were platform uptake (cumulative registrations, users sending at least one message) and repeat engagement (users meeting the pre-specified active-use threshold). Secondary outcomes were geographic and language distribution, message domain and intent, dosing-card activation, and explicit user feedback events. No accuracy, safety, or clinical-effectiveness outcomes were assessed.

### Patient and public involvement

This evaluation concerned a tool intended solely for healthcare professionals and researchers with a professional interest in IBD. No patients or members of the public were involved in the design, conduct, or reporting of this service evaluation.

### Governance and ethics

This work was conducted as a descriptive evaluation of a live digital service using aggregated, de-identified operational metrics generated during routine operation. Under Health Research Authority guidance, the work was treated as service evaluation rather than research. The platform operates in compliance with the UK General Data Protection Regulation and the Data Protection Act 2018.

Language and access equity were recognised as fairness considerations given multilingual use over a predominantly English-language corpus. No formal fairness or bias auditing was performed; evaluation of non-English performance is a stated future validation priority.

## Results

### Registrations, activity, and growth

During the first 6 months of deployment, ChatIBD registered 913 users (Figure 3A). The platform launched on 1 October 2025, and early dissemination through social media and an existing professional IBD network was reflected in the initial growth pattern, with 512 new registrations in October 2025. 684 (74.9%) users sent at least 1 message during the study period and 349 (38.2%) sent at least 3 messages and met the pre-specified definition of active use.

**Figure 3.**
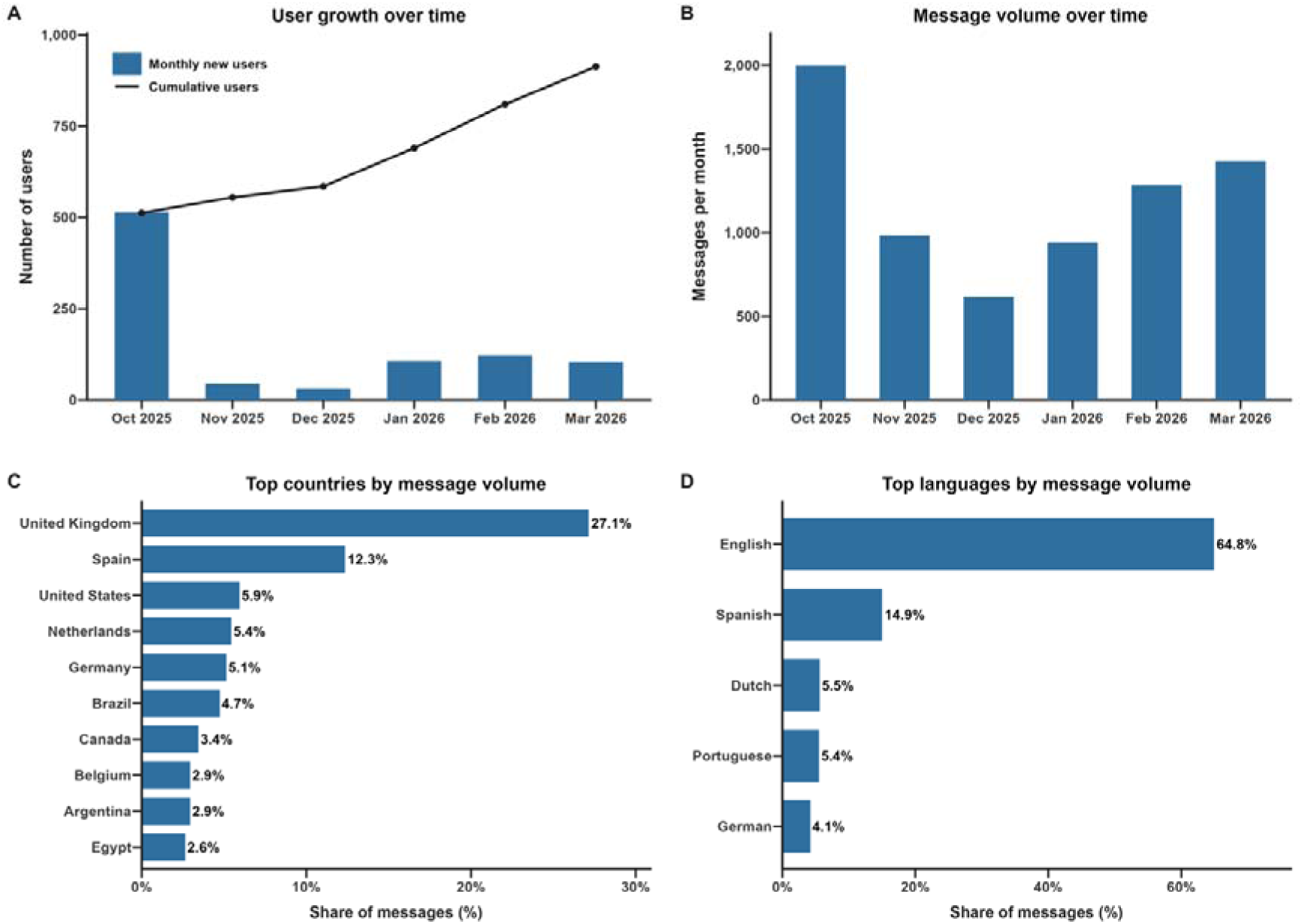
Early growth, message volume, and international uptake of ChatIBD during the first 6 months of live deployment. **A:** Monthly new registrations and cumulative registered users. **B:** Monthly message volume. **C:** Top 10 countries by share of total messages. **D:** Top 5 languages by share of total messages.

Across the full study period, the platform processed 7,222 user messages across 3,855 conversations. Median daily message volume was 35.5 (IQR 20 to 52). Median messages per conversation was 2 (IQR 2 to 4), consistent with use as a rapid reference tool rather than a prolonged conversational assistant (Figure 3B). Usage was concentrated on weekdays, which accounted for 6,145 messages (85.1%), compared with 1,077 (14.9%) on weekends. Activity was highest on Wednesdays and Thursdays, and lowest on Sundays.

### Geographic and language distribution

Message activity was recorded across 69 countries. The largest message volumes were from the United Kingdom (1,956 messages, 27.1%), Spain (887, 12.3%), the United States (428, 5.9%), the Netherlands (391, 5.4%), and Germany (368, 5.1%) (Figure 3C). 28 languages were recorded and English accounted for 4,681 messages (64.8%), followed by Spanish (1,078, 14.9%), Dutch (394, 5.5%), Portuguese (388, 5.4%), and German (293, 4.1%) (Figure 3D).

### Optional professional profile data

Among the 184 users with profile data, 115 (62.5%) identified as consultant or attending gastroenterologists, 26 (14.1%) as trainees or fellows, 19 (10.3%) as IBD nurse specialists, and 24 (13.0%) as allied health professionals.

### User feedback and flagged outputs

During the study period, 16 explicit user feedback events were recorded overall, comprising 15 positive ratings and 1 negative rating. Relative to total platform activity, feedback was infrequent. The one interaction explicitly flagged as a potential clinical error related to ambiguity in the response sentence, “Risankizumab is non-inferior to ustekinumab in anti-TNF-exposed, but not bio-naive, patients”, which risked implying inferiority in biologic-naive patients when the intended meaning was that direct comparative evidence in that subgroup was lacking. This was subsequently mitigated through changes to the production system. This case is presented as an example of the platform’s review workflow rather than as a formal safety outcome.

### Message domains and intents

Medication-related messages represented a substantial proportion of platform activity, with 3,364 messages (46.6%) classified within a pharmacotherapy domain (Table 2). The dosing card was triggered 1,332 times, reflecting the platform’s two-tier approach to medication questions. Exploratory LLM-based classification suggested that use also extended to education and score interpretation (737, 10.2%), infection and immunisation (548, 7.6%), reproduction and lactation (485, 6.7%), and surveillance and monitoring (473, 6.5%). At the intent level, guideline synthesis was the most frequent category (2,199, 30.4%), followed by definition or teaching messages (1,110, 15.4%), safety or contraindication messages (1,096, 15.2%), and treatment sequencing or algorithm questions (1,056, 14.6%).

**Table 2.** Exploratory LLM-based classification of user messages by domain and intent during the study period.

| Domain | Messages, n | % | Intent | Messages, n | % |
| --- | --- | --- | --- | --- | --- |
| Pharmacotherapy | 3,364 | 46.6 | Guideline synthesis | 2,199 | 30.4 |
| Education and score interpretation | 737 | 10.2 | Definition or teaching | 1,110 | 15.4 |
| Infection and immunisation | 548 | 7.6 | Safety or contraindication | 1,096 | 15.2 |
| Unclassified | 504 | 7.0 | Treatment sequencing or algorithm | 1,056 | 14.6 |
| Reproduction and lactation | 485 | 6.7 | Risk counselling | 708 | 9.8 |
| Surveillance and monitoring | 473 | 6.5 | Case triage or next step | 693 | 9.6 |
| Diagnostics and laboratory interpretation | 424 | 5.9 | Dose lookup | 582 | 8.1 |
| Procedures and perioperative care | 361 | 5.0 | Score table request | 110 | 1.5 |
| Comorbidities and special populations | 326 | 4.5 |  |  |  |

*Domain and intent labels were assigned using an exploratory large language model-based classifier (grok-4-fast-reasoning) as part of routine service monitoring and were not independently validated. Each user message was assigned a single primary domain. Messages could receive one or more intent labels.*

## Discussion

We report early uptake and repeat engagement with a specialty-specific retrieval-grounded clinical AI tool deployed with explicit safeguards, although usage alone does not establish safety, accuracy, or clinical effectiveness. The median of 2 messages per conversation is consistent with its intended use as a rapid specialist reference tool rather than a prolonged conversational assistant. International and multilingual use, despite the absence of formal health system deployment or large-scale promotion, suggests demand for faster navigation of specialist guidance. It may also indicate scope to widen access to predominantly English-language guideline content, although non-English performance requires formal evaluation.

These findings are consistent with growing interest in specialty-specific retrieval-grounded clinical AI tools.(10) Prior technical evaluations in gastroenterology suggest retrieval-augmented approaches may improve faithfulness and traceability compared with unconstrained generation.(11) ChatIBD adds a complementary real-world deployment perspective.

From a regulatory perspective, large language model-based clinical AI poses a challenge that is not yet fully resolved: validation is unlikely to be a one-off event.(12,13) Our experience suggests that safe live deployment depends on continuous monitoring, feedback capture, and the ability to review and respond quickly to problematic outputs.(14) This is especially relevant for systems built on foundation models that may be updated, deprecated, or replaced, and whose behaviour may also shift with changes to prompts, retrieval pipelines, or source content. In this setting, the central question is not only whether a system was validated before deployment, but how it should be monitored and revalidated over time.

Strengths of this evaluation include real-world multinational usage data, a per-message model and prompt audit trail supporting reconstruction of the exact configuration behind any response, governance-native design, and a deterministic structured dosing layer separated from free-text generation.

There are several important limitations. First, this was a descriptive analysis of early operational data from a self-selected user base. Similarly, the low volume of explicit user feedback should not be interpreted as evidence of a low error rate. Initial dissemination through social media and professional networks means early adopters are unlikely to be representative of the wider IBD workforce. Second, this study did not formally evaluate response accuracy, completeness, citation fidelity, consistency with source guidelines, or appropriateness in specific clinical scenarios. Third, no patient-level, workflow-level, or clinician decision-level outcomes were measured.

In addition, platform behaviour was not fully static during the observation period: the production model was updated from GPT-4.1 to GPT-5.1 and three source documents were added to the knowledge base. Flagged-response review was also conducted by a single author during this early phase without external adjudication. Exploratory domain and intent classifications were generated using an LLM-based classifier and should be interpreted as descriptive approximations rather than validated annotations.

Formal validation is in progress, with priorities including response accuracy, citation fidelity, medication-related performance, non-English behaviour, and impact on clinical workflow. More rigorous evaluation will be needed before systems of this kind can be relied upon more widely in practice.

## Conclusion

ChatIBD showed early international uptake and repeat use as a specialty-specific, guideline-grounded generative AI tool for inflammatory bowel disease, deployed with a set of practical operational safeguards that we describe in detail. These usage findings do not by themselves establish clinical accuracy, safety, or effectiveness. Formal validation is in progress.

## Declarations

### Data and code availability

Aggregated platform metrics used in this analysis are publicly available at https://github.com/shaunchuah/chatibd_platform_metrics. The platform source code is proprietary and constitutes intellectual property of the University of Glasgow.

### Author contributions

CSC conceived and designed the platform, led the technical implementation, performed the data analysis, and drafted the manuscript. BG led user engagement, dissemination, and community networking, and contributed to platform content curation. NP led the research and validation strategy and contributed to study design. All authors contributed to the selection of the guideline knowledge base, reviewed and revised the manuscript, and approved the final version.

### Tables, figures, and supplementary material

**Supplementary Table 1.** Source documents included in the ChatIBD knowledge base during the study period, with originating society or organisation, topic area, document title, and publication year.

| **Society(s)** | **Topic** | **Publication Name** | **Year** |
| --- | --- | --- | --- |
| ACG | Adult UC | ACG Clinical Guideline Update: Ulcerative Colitis in Adults | 2025 |
| ACG | Adult CD | ACG Clinical Guideline: Management of Crohn's Disease in Adults | 2025 |
| AGA | Adult CD | AGA Living Clinical Practice Guideline on the Pharmacologic Management of Moderate-to-Severe Crohn's Disease | 2025 |
| BSG | Malignancy and surveillance | British Society of Gastroenterology guidelines on colorectal surveillance in inflammatory bowel disease | 2025 |
| BSG | Adult UC and CD | British Society of Gastroenterology guidelines on inflammatory bowel disease in adults: 2025 | 2025 |
| ECCO | Diet | ECCO Consensus on Dietary Management of Inflammatory Bowel Disease | 2025 |
| ECCO, ESGAR, ESP, IBUS | Diagnosis and monitoring | ECCO-ESGAR-ESP-IBUS Guideline on Diagnostics and Monitoring of Patients with Inflammatory Bowel Disease: Part 1 | 2025 |
| ECCO, ESGAR, ESP, IBUS | Diagnosis and monitoring | ECCO-ESGAR-ESP-IBUS Guideline on Diagnostics and Monitoring of Patients with Inflammatory Bowel Disease: Part 2 | 2025 |
| ECCO, ESPGHAN | Paediatric UC | ECCO/ESPGHAN Management of paediatric ulcerative colitis, part 1 | 2025 |
| ECCO, ESPGHAN | Paediatric UC | ECCO/ESPGHAN Management of paediatric ulcerative colitis, part 2 | 2025 |
| Global Consensus Consortium | Fertility and pregnancy | Global Consensus Statement on the Management of Pregnancy in Inflammatory Bowel Disease | 2025 |
| UKHSA | Vaccinations | Green book chapter: Shingles (herpes zoster) | 2025 |
| UKHSA | Vaccinations | Green book chapter: Varicella | 2025 |
| Johnson & Johnson Safety Update | Medication safety | STELARA - Occurrence of Posterior Reversible Encephalopathy Syndrome (PRES) | 2025 |
| GETECCU | Adult UC and CD | Spanish Working Group in Crohn's Disease and Ulcerative Colitis (GETECCU) position paper on cardiovascular disease in patients with inflammatory bowel disease | 2025 |
| AGA | Pouchitis | AGA Clinical Practice Guideline on the Management of Pouchitis and Inflammatory Pouch Disorders | 2024 |
| AGA | Adult UC | AGA Living Clinical Practice Guideline on Pharmacological Management of Moderate-to-Severe Ulcerative Colitis | 2024 |
| BSG | Endoscopy | British Society of Gastroenterology guidelines on sedation in gastrointestinal endoscopy | 2024 |
| ECCO | Pathology | Definitions of Histological Abnormalities in Inflammatory Bowel Disease: an ECCO Position Paper | 2024 |
| ECCO | Adult CD | ECCO Guidelines on Therapeutics in Crohn's Disease: Medical Treatment | 2024 |
| ECCO | Adult CD | ECCO Guidelines on Therapeutics in Crohn's Disease: Surgical Treatment | 2024 |
| AGA | Stoma management | AGA Clinical Practice Update on Management of Ostomies: Commentary | 2023 |
| ECCO | Extra intestinal manifestations | ECCO Guidelines on Extraintestinal Manifestations in Inflammatory Bowel Disease | 2023 |
| ECCO | Malignancy and surveillance | ECCO Guidelines on Inflammatory Bowel Disease and Malignancies | 2023 |
| ECCO | Fertility and pregnancy | European Crohn's and Colitis Guidelines on Sexuality, Fertility, Pregnancy, and Lactation | 2023 |
| ECCO | Adult CD | Results of the Eighth Scientific Workshop of ECCO: Prevention and Treatment of Postoperative Recurrence in Patients With Crohn's Disease Undergoing an Ileocolonic Resection With Ileocolonic Anastomosis | 2023 |
| AGA | Short bowel syndrome | AGA Clinical Practice Update on Management of Short Bowel Syndrome: Expert Review | 2022 |
| ECCO | Adult UC | ECCO Guidelines on Therapeutics in Ulcerative Colitis: Medical Treatment | 2022 |
| GETECCU | Adult CD | Position Statement. Recommendations of the Spanish Group on Crohn's Disease and Ulcerative Colitis (GETECCU) on the treatment of strictures in Crohn's disease | 2022 |
| GETECCU | Vaccinations | Recommendations of the Spanish Group on Crohn's Disease and Ulcerative Colitis on the importance, screening and vaccination in inflammatory bowel disease patients | 2022 |
| BSG, ESGE | Endoscopy | Endoscopy in patients on antiplatelet or anticoagulant therapy: BSG and ESGE guideline update | 2021 |
| ECCO, ESPGHAN | Paediatric CD | The Medical Management of Paediatric Crohn's Disease: an ECCO-ESPGHAN Guideline Update | 2021 |
| BSG, ACGBI, PHE | Malignancy and surveillance | BSG/ACGBI/PHE post- polypectomy and post- colorectal cancer resection surveillance guidelines | 2020 |
| UEG | Microscopic colitis | European guidelines on microscopic colitis: United European Gastroenterology and European Microscopic Colitis Group statements and recommendations | 2020 |
| AGA | Malignancy and surveillance | AGA Clinical Practice Update on Surveillance for Hepatobiliary Cancers in Patients With Primary Sclerosing Cholangitis: Expert Review | 2019 |

**Abbreviations:** ACG, American College of Gastroenterology; ACGBI, Association of Coloproctology of Great Britain and Ireland; AGA, American Gastroenterological Association; BSG, British Society of Gastroenterology; ECCO, European Crohn's and Colitis Organisation; ESGAR, European Society of Gastrointestinal and Abdominal Radiology; ESGE, European Society of Gastrointestinal Endoscopy; ESP, European Society of Pathology; ESPGHAN, European Society for Paediatric Gastroenterology, Hepatology and Nutrition; GETECCU, Grupo Español de Trabajo en Enfermedad de Crohn y Colitis Ulcerosa; IBUS, International Bowel Ultrasound Group; PHE, Public Health England; UKHSA, UK Health Security Agency; UEG, United European Gastroenterology

**Supplementary Table 2.** DECIDE-AI (2022) reporting checklist for ChatIBD.

| Item | Theme | Recommendation | Status |
| --- | --- | --- | --- |
| Title and abstract |  |  |  |
| 1 | Title | Identify the study as early clinical evaluation of an AI/ML decision-support system, specifying the problem addressed. | Yes |
| I | Abstract | Provide a structured summary (intended use, algorithm type, setting, users, outcomes, results, conclusions). | Yes |
| Introduction |  |  |  |
| 2a | Intended use | Describe the targeted condition(s), problem(s), current standard practice, and intended patient population. | Yes |
| 2b | Intended use | Describe the intended users, planned integration in the care pathway, and intended impact. | Yes - standalone tool, not integrated into any clinical workflow. |
| II | Objectives | State the study objectives. | Yes |
| Methods |  |  |  |
| III | Research governance | Reference any protocol, registration number, and ethics approval. | Yes - HRA tool confirmed service evaluation; registration/protocol not required. |
| 3a | Participants | Describe patient recruitment and inclusion/exclusion at patient and data level. | N/A - no patients or patient-level data. |
| 3b | Participants | Describe user recruitment, inclusion/exclusion, and how user numbers were decided. | Yes - open public deployment; self-selected users, no pre-specified sample size. |
| 3c | Participants | Describe steps to familiarise users with the system, including training. | Yes - on-screen instructions and disclaimers; no formal training (open-access tool). |
| 4a | AI system | Describe the system, version, and algorithm type; training population and preclinical performance. | Partial - versions and RAG architecture described; vendor foundation model, no preclinical study. |
| 4b | AI system | Identify inputs; describe acquisition, pre-processing, and handling of missing/low-quality data. | Yes |
| 4c | AI system | Describe the system outputs and how they were presented to users. | Yes - cited responses and dosing cards (Figure 2). |
| 5a | Implementation | Describe the settings in which the system was evaluated. | Yes - live, open-access, international web deployment. |
| 5b | Implementation | Describe the workflow/care pathway, timing of use, and how the decision was reached and by whom. | Yes - standalone use at user discretion; user retains all decisions. |
| IV | Outcomes | Specify the primary and secondary outcomes measured. | Yes - pre-specified descriptive usage outcomes; no clinical-performance outcomes. |
| 6a | Safety and errors | Describe how significant errors/malfunctions were defined and identified. | Yes - user flagging and clinician review. |
| 6b | Safety and errors | Describe how risks to patient safety/harm were identified, analysed, and minimised. | Yes - layered safeguards (Table 1) |
| 7 | Human factors | Describe human-factors tools/methods/frameworks, use cases, and users involved. | N/A - no formal human-factors evaluation conducted. |
| V | Analysis | Describe the statistical methods and any pre-specified additional analyses. | Yes - descriptive only (medians/IQR, counts); no inferential testing. |
| 8 | Ethics | Describe any methodologies used for an ethics-related goal (e.g. algorithmic fairness). | Yes - governance, data protection, and PII safeguards |
| VI | Patient involvement | State how patients were involved in the research question, design, and conduct. | Yes - none; HCP-facing tool. |
| Results |  |  |  |
| 9a | Participants | Report baseline patient characteristics and input-data missingness. | N/A - no patients. |
| 9b | Participants | Report baseline characteristics of the users included. | Yes - geography, language, optional professional background (engaged subset). |
| 10a | Implementation | Report user exposure, number of uses, and adherence to intended implementation. | Yes |
| 10b | Implementation | Report significant changes to the workflow/care pathway caused by the system. | N/A - standalone tool; no care-pathway changes measured. |
| VII | Main results | Report on pre-specified outcomes, including any comparison group. | Yes - single-arm descriptive; no comparison group. |
| VIII | Subgroups analysis | Report differences in main outcomes by pre-specified subgroups. | Yes - descriptive breakdowns only; no inferential subgroup comparison. |
| 11 | Modifications | Report changes to the system during the study, with timing, rationale, and outcome effects. | Yes - GPT-4.1 to GPT-5.1 (18 Nov 2025) and three documents added; outcome effect not formally measured. |
| 12 | Human-computer agreement | Report user agreement with, and variation from, system recommendations. | N/A - user agreement/decision change not captured. |
| 13a | Safety and errors | List significant errors/malfunctions with rate, causes, correctability, and impact. | Partial - one flagged response reviewed and mitigated; no systematic error-rate quantification. |
| 13b | Safety and errors | Report risks to patient safety or observed harm (including indirect harm). | Yes - no harm identified; patient-level harm not assessable (no patient data). |
| 14a | Human factors | Report the usability evaluation per recognised standards/frameworks. | N/A - usability not formally evaluated. |
| 14b | Human factors | Report the user learning-curve evaluation. | N/A - learning curves not evaluated. |
| Discussion |  |  |  |
| 15 | Support for intended use | Discuss whether results support the intended use in clinical settings. | Yes - usage supports feasibility; does not establish effectiveness. |
| 16 | Safety and errors | Discuss what results indicate about the safety profile, errors, harm, and mitigation. | Yes - safety not established; continuous monitoring and revalidation discussed. |
| IX | Strengths and limitations | Discuss the strengths and limitations of the study. | Yes |

| Statements |  |  |  |
| --- | --- | --- | --- |
| 17 | Data availability | Disclose if and how data and relevant code are available. | Yes |
| X | Conflicts of interest | Disclose conflicts, funding source, funder role, and per-author interests. | Yes |

## Data Availability

Aggregated platform metrics used in this analysis are available from the corresponding author on reasonable request.

## References

1. Kaplan GG. The global burden of inflammatory bowel disease: from 2025 to 2045. Nat Rev Gastroenterol Hepatol. 2025 Oct;22(10):708–20. doi:10.1038/s41575-025-01097-1

2. Caballero Mateos AM, Cañadas de la Fuente GA, Gros B. Paradigm Shift in Inflammatory Bowel Disease Management: Precision Medicine, Artificial Intelligence, and Emerging Therapies. J Clin Med. 2025 Feb 25;14(5):1536. doi:10.3390/jcm14051536 PubMed PMID: 40095460; PubMed Central PMCID: PMC11899940.

3. Rao AS, Esmail KP, Lee RS, Jiang S, Arraiza Carlo B, Gill J, et al. Large Language Model Performance and Clinical Reasoning Tasks. JAMA Netw Open. 2026 Apr 13;9(4):e264003. doi:10.1001/jamanetworkopen.2026.4003

4. Hager P, Jungmann F, Holland R, Bhagat K, Hubrecht I, Knauer M, et al. Evaluation and mitigation of the limitations of large language models in clinical decision-making. Nat Med. 2024 Sep;30(9):2613–22. doi:10.1038/s41591-024-03097-1

5. Bélisle-Pipon JC. Why we need to be careful with LLMs in medicine. Front Med. 2024 Dec 4;11. doi:10.3389/fmed.2024.1495582

6. Ong JCL, Chang SYH, William W, Butte AJ, Shah NH, Chew LST, et al. Ethical and regulatory challenges of large language models in medicine. The Lancet Digital Health. 2024 Jun 1;6(6):e428–32. doi:10.1016/S2589-7500(24)00061-X PubMed PMID: 38658283.

7. Gargari OK, Habibi G. Enhancing medical AI with retrieval-augmented generation: A mini narrative review. Digit Health. 2025;11:20552076251337177. doi:10.1177/20552076251337177 PubMed PMID: 40343063; PubMed Central PMCID: PMC12059965.

8. Amugongo LM, Mascheroni P, Brooks S, Doering S, Seidel J. Retrieval augmented generation for large language models in healthcare: A systematic review. PLOS Digital Health. 2025 Jun 11;4(6):e0000877. doi:10.1371/journal.pdig.0000877

9. Vasey B, Nagendran M, Campbell B, Clifton DA, Collins GS, Denaxas S, et al. Reporting guideline for the early-stage clinical evaluation of decision support systems driven by artificial intelligence: DECIDE-AI. Nat Med. 2022 May;28(5):924–33. doi:10.1038/s41591-022-01772-9

10. Ke YH, Jin L, Elangovan K, Abdullah HR, Liu N, Sia ATH, et al. Retrieval augmented generation for 10 large language models and its generalizability in assessing medical fitness. npj Digit Med. 2025 Apr 5;8(1):187. doi:10.1038/s41746-025-01519-z

11. Zhou Q, Liu C, Duan Y, Sun K, Li Y, Kan H, et al. GastroBot: a Chinese gastrointestinal disease chatbot based on the retrieval-augmented generation. Front Med (Lausanne). 2024;11:1392555. doi:10.3389/fmed.2024.1392555 PubMed PMID: 38841582; PubMed Central PMCID: PMC11150590.

12. Rosenthal JT, Beecy A, Sabuncu MR. Rethinking clinical trials for medical AI with dynamic deployments of adaptive systems. npj Digit Med. 2025 May 6;8(1):252. doi:10.1038/s41746-025-01674-3

13. Freyer O, Wiest IC, Kather JN, Gilbert S. A future role for health applications of large language models depends on regulators enforcing safety standards. The Lancet Digital Health. 2024 Sep 1;6(9):e662–72. doi:10.1016/S2589-7500(24)00124-9 PubMed PMID: 39179311.

14. Asgari E, Montaña-Brown N, Dubois M, Khalil S, Balloch J, Yeung JA, et al. A framework to assess clinical safety and hallucination rates of LLMs for medical text summarisation. npj Digit Med. 2025 May 13;8(1):274. doi:10.1038/s41746-025-01670-7

